# Myocarditis in a pediatric emergency department: a 10-year, single-center retrospective study

**DOI:** 10.1101/2023.05.25.23290562

**Authors:** Doyoung Jung

**Affiliations:** Division of Pediatric Emergency, Severance Children Hospital, Department of Pediatrics, Yonsei University College of Medicine, Seoul, Republic of Korea

## Abstract

**Background:** Pediatric myocarditis is caused by myocardial inflammation that can manifest a wide range of clinical courses, from mild symptoms to death, in the pediatric emergency department. This study aimed to identify risk factors or parameters to predict the clinical course of pediatric myocarditis.

**Methods:** This retrospective study included 46 patients who visited the pediatric emergency room at Severance Hospital. The patients were classified into spontaneous recovery (n=42) and extracorporeal membrane oxygenation (ECMO, n=4) groups, and their baseline characteristics, laboratory studies, and echocardiography findings were compared.

**Results:** Factors showing significant differences between the two groups included increased levels of inflammatory markers (delta neutrophil index, erythrocyte sedimentation rate, and C-reactive protein), aspartate aminotransferase (AST), and cardiac enzymes (creatine kinase-MB, N-terminal pro-B-type natriuretic peptide, and troponin-T) (p<0.001).

**Conclusions:** This study identified risk factors for fulminant myocarditis in pediatric patients with myocarditis who visited the emergency room, including increased levels of inflammatory markers, AST, and cardiac enzymes. Close monitoring and preparation for ECMO may be necessary if these risk factors are observed. These findings will be helpful for the early diagnosis and appropriate treatment of pediatric myocarditis.

## Introduction

Myocarditis is a rare disease that occurs due to inflammation of heart muscle cells, mainly in pediatric patients.1-4 The reported prevalence is 3.5–5%, and most cases have a mild course.5 However, myocarditis is a life-threatening condition in 0.1–0.6% of cases.6 Although patients present to the pediatric emergency department with a wide range of symptoms, diagnosing myocarditis is not easy.7-9 According to one review, myocarditis in the emergency department was reported in 0.5 cases per 10,000 patients.10 Additionally, there are no definitive clinical symptoms, making it a difficult disease to diagnose.11

Laboratory tests,12-14 chest X-rays, EKG,15-18 and echocardiography can be used to aid in diagnosis, but confirmation requires invasive methods such as endomyocardial biopsy or cardiac magnetic resonance imaging (MRI),19 which can be difficult to perform or require too much time to perform in unstable patients. Therefore, there are no clear criteria for diagnosing myocarditis or identifying risk factors for fulminant myocarditis.

This study aimed to retrospectively analyze the characteristics and clinical course of pediatric patients with myocarditis who visited the emergency department to identify factors that may influence prognosis and aid in appropriate treatment.

## Methods

This study included 46 patients aged <18 years who were diagnosed with myocarditis during their visits to the Pediatric Emergency Department of Severance Hospital between January 2003 and December 2022. All patients underwent blood tests, chest X-ray (CXR), and echocardiography. Patients with elevated cardiac enzyme levels or suspected myocarditis based on EKG abnormalities or echocardiography were included in the present study. Blood tests including complete blood count (CBC); blood chemistry; and measurement of creatine kinase (CK), CK-MB, N-terminal pro-B-type natriuretic peptide (NT-proBNP), and troponin-T levels were performed at the time of emergency room visits. Electrocardiograms (EKGs) were performed at the time of emergency room visits and were followed up according to symptoms to check for abnormalities. Chest radiography was also performed during emergency room visits, mostly in posterior-anterior (PA) views; however, in some younger or critically ill patients, anterior-posterior (AP) views were used.

Echocardiography was performed in patients with typical cardiac symptoms (chest pain, dyspnea, and palpitations) and elevated cardiac enzyme levels. Transthoracic echocardiography was performed, and the parasternal long-axis, short-axis, and apical four-chamber views were routinely observed. Left ventricular ejection fraction (LVEF), tissue Doppler, and interventricular septum (IVS) thickness were measured in all patients. In this study, patients were classified into two groups based on disease severity: spontaneous recovery and ECMO. Factors influencing severity were statistically analyzed.

The study was conducted according to the research protocol approved by the Institutional Review Board of Severance Hospital (IRB 4-2023-0117) and the Declaration of Helsinki.

A descriptive analysis of nominal variables was conducted using numbers and percentages, and univariate analysis was performed using the chi-squared test. To analyze factors that influenced poor prognosis, multivariate logistic regression analysis was performed, including variables with P < 0.05 in the univariate analysis as input variables. Odds ratios (ORs) and 95% confidence intervals (CIs) were calculated, and statistical significance was defined as P < 0.05. IBM SPSS Statistics for Windows, version 26.0 (IBM Corp., Armonk, NY, USA) was used for statistical analysis.

## Results

A total of 46 patients with suspected myocarditis were admitted to the Pediatric Emergency Department of Severance Hospital. At the time of admission, the average age was 7.36 years, with a bimodal distribution in the 0–5 and 10–17-year age groups. The average weight was 31 kg and the mean length of stay was 2.15 ± 6.17 days. Four patients underwent ECMO cannulation, among which two patients died.

The patients’ medical histories revealed that three patients (6.5%) had previously been diagnosed with myocarditis, two patients (4.3%) had Kawasaki disease, two patients (4.3%) had undergone surgery for other congenital heart diseases (ventricular septal defect [VSD], total anomalous pulmonary venous return [TAPVR]), and one patient (2.1%) had received chemotherapy for Burkitt lymphoma. The most common presenting symptom was dyspnea (10 patients, 21.7%), followed by fever (eight patients, 17.4%), gastrointestinal symptoms (vomiting and abdominal pain) (six patients, 13.0%), chest pain (six patients, 13.0%), syncope (four patients, 8.7%), and dizziness (four patients, 8.7%). Other reported symptoms included palpitations (three patients, 6.5%) and chest discomfort, auria, paleness, and drug intoxication in one case each.

Cardiomegaly was observed in five patients (10.8%) on chest radiography. The most common EKG abnormality was normal sinus rhythm (21 patients, 45.7%), followed by sinus tachycardia (12 patients, 26.0%), and ST elevation (two patients, 4.3%). Other abnormalities included prolonged QT and atrial tachycardia with PVC (premature ventricular contraction), RBBB (right bundle branch block), and left ventricular hypertrophy (LVH) in 1 case each (Table 1).

**Table 1.**
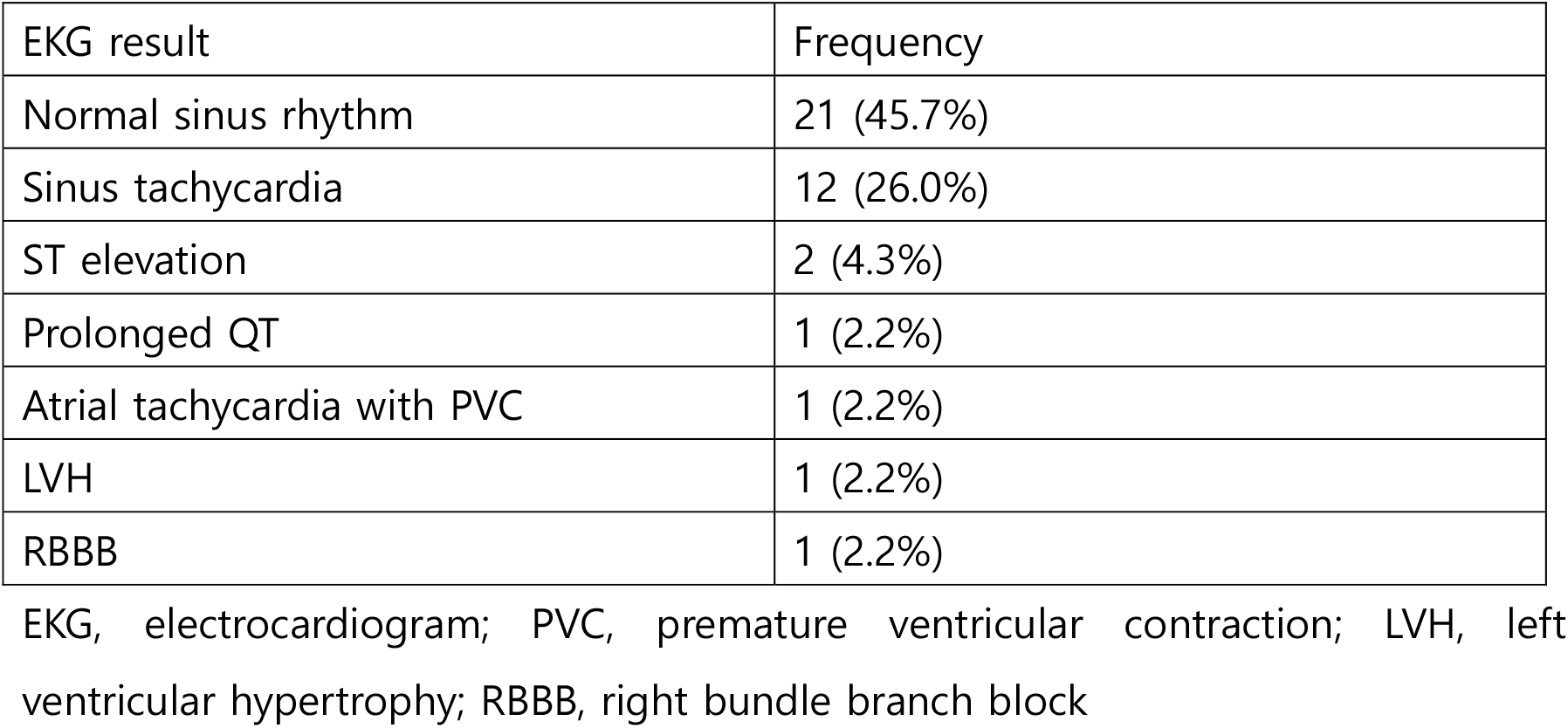
Electrocardiogram Findings at Presentation

Laboratory studies showed a mean WBC count of 9416.96 ± 4178.76/μl, delta neutrophil index (DNI) of 0.43 ± 1.15, erythrocyte sedimentation rate (ESR) of 7.13 ± 10.08 mm/h, and C-reactive protein (CRP) concentration of 6.31 ± 13.0 mg/L. The mean aspartate aminotransferase (AST) level was 88.61 ± 369.469 (U/L), while the concentrations of cardiac enzymes such as CK-MB, NT-proBNP, and troponin-T were 8.49 ± 28.28 IU/L, 1762.0 ± 6119.44 pg/mL, and 30.07 ± 90.03 ng/mL, respectively.

Echocardiography showed a mean LVEF of 61.43 ± 15.60%, E/A of 1.58 ± 0.59, RV TDI s’ of 10.59 ± 2.68 cm/s, and IVS thickness of 8.71 ± 2.99 mm.

A comparison between the spontaneous recovery (n=42) and the ECMO (n=4) groups showed no significant differences in age, weight, or other demographic characteristics. However, some laboratory studies differed significantly, including DNI, ESR, CRP, and AST (p<0.001), as well as cardiac enzymes such as CK-MB, NT-proBNP, and troponin T (p<0.001). Echocardiography features did not differ significantly between the two groups (Table 2).

**Table 2.**
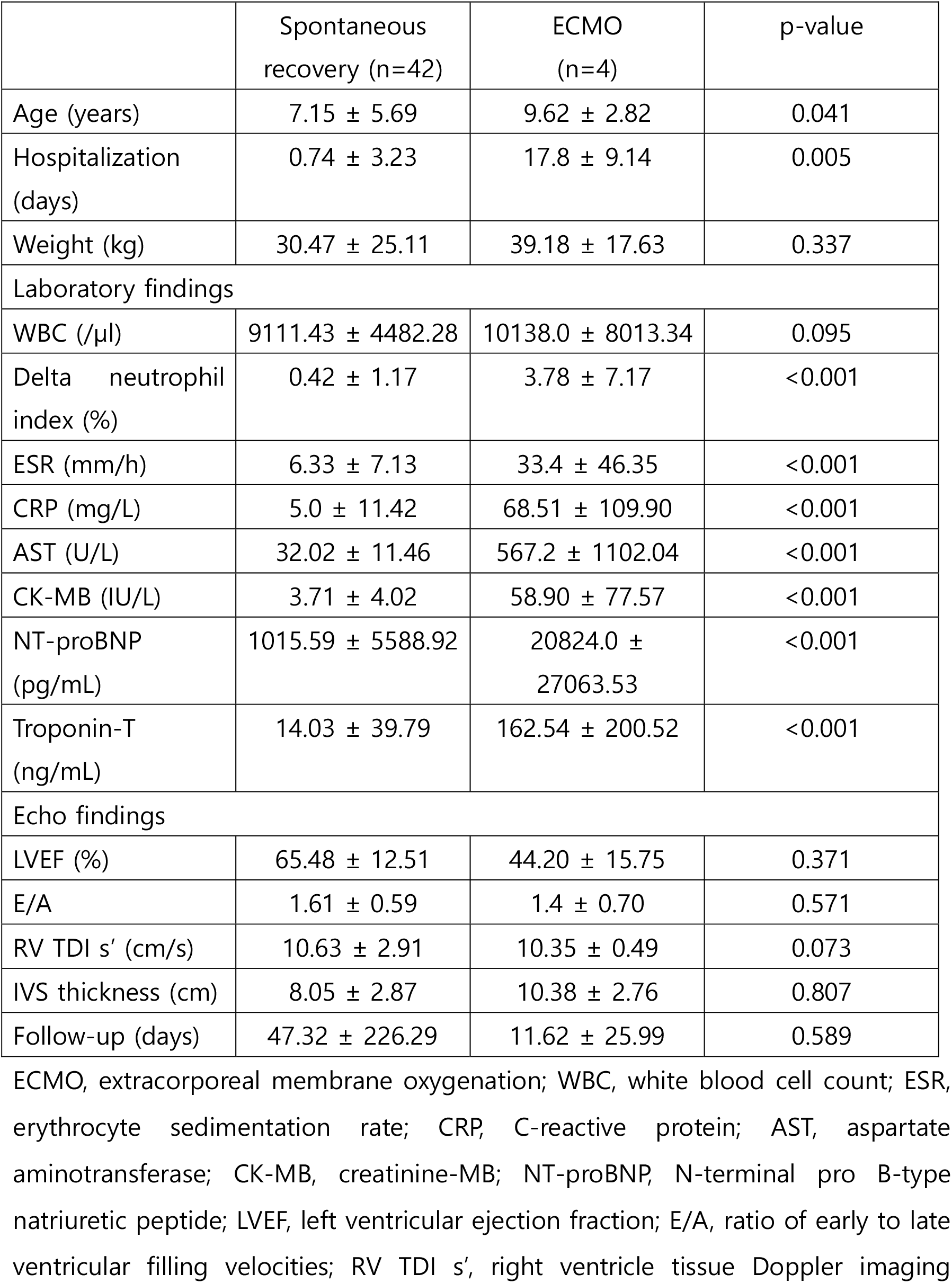
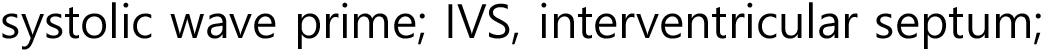
Baseline Patient Characteristics

|  | Spontaneous recovery (n=42) | ECMO (n=4) | p-value |
| --- | --- | --- | --- |
| Age (years) | 7.15 ± 5.69 | 9.62 ± 2.82 | 0.041 |
| Hospitalization (days) | 0.74 ± 3.23 | 17.8 ± 9.14 | 0.005 |
| Weight (kg) | 30.47 ± 25.11 | 39.18 ± 17.63 | 0.337 |
| Laboratory findings |  |  |  |
| WBC (/μl) | 9111.43 ± 4482.28 | 10138.0 ± 8013.34 | 0.095 |
| Delta neutrophil index (%) | 0.42 ± 1.17 | 3.78 ± 7.17 | <0.001 |
| ESR (mm/h) | 6.33 ± 7.13 | 33.4 ± 46.35 | <0.001 |
| CRP (mg/L) | 5.0 ± 11.42 | 68.51 ± 109.90 | <0.001 |
| AST (U/L) | 32.02 ± 11.46 | 567.2 ± 1102.04 | <0.001 |
| CK-MB (IU/L) | 3.71 ± 4.02 | 58.90 ± 77.57 | <0.001 |
| NT-proBNP (pg/mL) | 1015.59 ± 5588.92 | 20824.0 ± 27063.53 | <0.001 |
| Troponin-T (ng/mL) | 14.03 ± 39.79 | 162.54 ± 200.52 | <0.001 |
| Echo findings |  |  |  |
| LVEF (%) | 65.48 ± 12.51 | 44.20 ± 15.75 | 0.371 |
| E/A | 1.61 ± 0.59 | 1.4 ± 0.70 | 0.571 |
| RV TDI s' (cm/s) | 10.63 ± 2.91 | 10.35 ± 0.49 | 0.073 |
| IVS thickness (cm) | 8.05 ± 2.87 | 10.38 ± 2.76 | 0.807 |
| Follow-up (days) | 47.32 ± 226.29 | 11.62 ± 25.99 | 0.589 |

ECMO, extracorporeal membrane oxygenation; WBC, white blood cell count; ESR, erythrocyte sedimentation rate; CRP, C-reactive protein; AST, aspartate aminotransferase; CK-MB, creatinine-MB; NT-proBNP, N-terminal pro B-type natriuretic peptide; LVEF, left ventricular ejection fraction; E/A, ratio of early to late ventricular filling velocities; RV TDI s', right ventricle tissue Doppler imaging
systolic wave prime; IVS, interventricular septum;

## Discussion

We retrospectively examined myocarditis in 46 patients who visited the pediatric emergency department. The prevalence followed a bimodal pattern, which is similar to the findings of other studies. Male sex was predominant, which is consistent with the generally known sex ratio.20, 21 EKG changes were mainly characterized by sinus tachycardia, in contrast to other studies where ventricular tachycardia was relatively common.16 Although complete AV block has been reported,17 it was not observed in the present study. This difference may be attributed to the location of myocardial involvement, as VT may appear as a precursor symptom of heart failure. Atrial tachycardia and ST elevation have also been reported.

AST levels were significantly higher in the ECMO group, consistent with previous.7, 9 AST elevation may be more likely to occur in patients presenting deterioration that leads to hepatic congestion, although cases have been reported in which AST levels did not increase with myocarditis progression.20 In the case of fulminant myocarditis requiring ECMO, heart failure progresses to a severe degree, which is likely the reason for significant AST elevation.

DNI is an indicator that can increase during acute infectious diseases.22 In this study, the DNI was also an indicator of fulminant myocarditis. Because it is mainly an indicator of sepsis, it was expected that it would not be greatly reflected in cases of localized infection, such as myocarditis; however, many cases of fulminant myocarditis showed increased DNI. This may occur due to changes in neutrophils resulting from inflammation, leading to systemic malnutrition; however, the exact cascade remains unclear.23 Further studies are required to clarify these points.

In this study, mixed cases of decreased and maintained LVEF were observed using echocardiography. Patients with classic myocarditis typically show decreased LVEF, normal ventricular cavity size, and septal thickening.24 However, in this study, clear decreases in LVEF were not observed, as cases with initial maintenance tended to worsen quickly in the emergency room. Two patients experienced immediate deterioration in the emergency room; these patients received ECMO and were transferred to the intensive care unit (ICU). The other two patients who received ECMO had relatively normal LVEF initially, but clear septal thickening was observed in the emergency room; thus, their conditions were carefully monitored. They subsequently deteriorated during hospitalization, and ECMO cannulation was performed. Therefore, even if LVEF is relatively maintained, septal thickening or regional wall motion abnormalities suggest a high possibility of rapid deterioration, thus requiring more careful monitoring.

Although the diagnosis of myocarditis was unclear in other studies, they have suggested that myocarditis can be suspected based on increased cardiac enzyme levels, ECG changes, and abnormal function on echocardiography or cardiac MRI (CMRI) due to myocardial damage.10, 25-27 The present study did not compare the myocarditis patient group to a normal control group and did not provide diagnostic criteria. More specific methods or the use of scoring are needed to improve the accuracy of the diagnosis.

In this study, the DNI; ESR; and CRP, CK-MB, NT-proBNP, and troponin-T levels were significantly higher in the ECMO group and were risk factors for fulminant myocarditis. Other studies have reported hypotension, elevated troponin-i/BPN levels, and decreased LVEF as indicators of fulminant myocarditis.28 Since the initial examinations were conducted in the emergency room in this study, the decrease in LVEF was not clearly visible. Increased levels of inflammatory markers and cardiac enzymes are indicative of fulminant myocarditis, while early changes in echocardiographic findings may not be apparent. Since the accuracy of echocardiography can vary depending on the examiner’s skill and this study had different examiners over a long study period, differences in echocardiographic parameters due to differences in skill levels may also explain the lack of significant differences in these parameters.

This study had several limitations. First, it was a retrospective study; therefore, there may have been missing information in the medical records or treatment. Second, the patient group was not compared with a normal control group; therefore, the differences between myocarditis and normal were not assessed. Finally, the small number of patients may have led to lower reliability of the findings.

## Conclusion

Pediatric myocarditis can lead to death in the emergency department. Cases of fulminant myocarditis, which can lead to ECMO cannulation, often show increased levels of inflammatory markers (ESR, CRP, and DNI), evidence of heart failure (increased AST and NT-proBNP levels), and increased levels of cardiac markers. In cases with these risk factors, careful monitoring and preparation for ECMO may be necessary. Further multicenter studies are needed to evaluate additional risk factors for a more definitive assessment.

## Data Availability

All data referred to in the manuscrip are available.

## Acknowledgements

We would like to thank Editage (www.editage.co.kr) for English language editing.

## References

1. Cooper Jr LT. Myocarditis. New England Journal of Medicine, 2009;15: 1526–1538. doi: 10.1056/NEJMra0800028

2. May, Lindsay J, Patton David J, Fruitman Deborah S. The evolving approach to paediatric myocarditis: a review of the current literature. Cardiology in the Young, 2011;21.3: 241–251. doi: 10.1017/S1047951110001964.

3. Caforio, Alida LP, Pankuweit, S, Arbustini E, Basso C, Gimeno-Blanes, J., et al. Current state of knowledge on aetiology, diagnosis, management, and therapy of myocarditis: a position statement of the European Society of Cardiology Working Group on Myocardial and Pericardial Diseases. European heart journal, 2013; 34.33: 2636–2648. doi: 10.1093/eurheartj/eht210.

4. Pettit, Mark A., Koyfman Alex, Foran Mark. Myocarditis. Pediatric emergency care, 2014;30.11: 832–835. doi: 10.1097/PEC.0000000000000272.

5. Schultheiss, Heinz-Peter, Kühl U, Cooper LT. The management of myocarditis. European heart journal, 2011;32.21: 2616–2625. doi: 10.1093/eurheartj/ehr165.

6. Levine, Marla C, Klugman, Darren, Teach Stephen J. Update on myocarditis in children. Current Opinion in Pediatrics, 2010;22.3: 278–283. doi: 10.1097/MOP.0b013e32833924d2.

7. Chang, Yi-Jung, et al. Myocarditis presenting as gastritis in children. Pediatric emergency care, 2006;22.6: 439–440. doi: 10.1097/01.pec.0000221346.64991.e7.

8. Shu-Ling, Chong, Bautista D, Chen CK, Su-Yin AA. Diagnostic evaluation of pediatric myocarditis in the emergency department: a 10-year case series in the Asian population. Pediatric Emergency Care, 2013;29.3: 346–351. doi: 10.1097/PEC.0b013e3182852f86.

9. Freedman, Stephen, Haladyn JK, Floh A, Kirsh JA, Taylor G, Thull-Freedman J. Pediatric myocarditis: emergency department clinical findings and diagnostic evaluation. Pediatrics, 2007; 120.6: 1278–1285. doi: 10.1542/peds.2007-1073.

10. Sagar S, Sandeep L, Peter P, Cooper LT. Myocarditis. The Lancet, 2012;379.9817: 738–747. doi: 10.1016/S0140-6736(11)60648-X.

11. Law YM, Lal AK, Chen S, Čiháková D, Cooper Jr LT, Deshpande S, et al. Diagnosis and management of myocarditis in children: a scientific statement from the American Heart Association. Circulation, 2021;144.6: e123–e135. doi: 10.1161/CIR.0000000000001001.

12. Smith SC, Ladenson JH, Mason, JW, Jaffe AS. Elevations of cardiac troponin I associated with myocarditis: experimental and clinical correlates. Circulation, 1997;95.1: 163–168. Doi: 10.1161/01.CIR.95.1.163

13. Lippi G, Salvagno GL, Guidi GC. Cardiac troponins in pediatric myocarditis. Pediatrics, 2008;121.864: 2008–0031. doi: 10.1542/peds.2008-0031.

14. Lauer, B, Niederau C, Kühl U, Schannwell M, Pauschinger M, Strauer BE, et al. Cardiac troponin T in patients with clinically suspected myocarditis. Journal of the American College of Cardiology, 1997;30.5: 1354–1359. doi: 10.1016/s0735-1097(97)00317-3.

15. Pompa AG, Beerman LB, Feingold B, Arora G. Electrocardiogram changes in pediatric patients with myocarditis. The American Journal of Emergency Medicine, 2022;59: 49–53. doi: 10.1016/j.ajem.2022.06.027.

16. Cartoski MJ, Nikolov, PP, Prakosa A., Boyle PM, Spevak PJ, Trayanova, NA. Computational identification of ventricular arrhythmia risk in pediatric myocarditis. Pediatric cardiology, 2019;40: 857–864. doi: 10.1007/s00246-019-02082-7.

17. Ichikawa R, Sumitomo N, Komori A, Abe Y, Nakamura T, Fukuhara J, et al. The follow-up evaluation of electrocardiogram and arrhythmias in children with fluminant myocarditis. Circulation Journal, 2011;75.4: 932–938. doi: 10.1253/circj.cj-10-0918.

18. Miyake CY, Teele SA, Chen L, Motonaga KS, Dubin AM, Balasubramanian S, et al. In-hospital arrhythmia development and outcomes in pediatric patients with acute myocarditis. The American journal of cardiology, 2014;113.3: 535–540. doi: 10.1016/j.amjcard.2013.10.021.

19. Friedrich MG, Sechtem U, Schulz-Menger J, Holmvang G, Alakija P, Cooper LT, et al. Cardiovascular magnetic resonance in myocarditis: A JACC White Paper. Journal of the American College of Cardiology, 2009;53.17: 1475–1487. doi: 10.1016/j.jacc.2009.02.007.

20. Ghelani SJ, Spaeder MC, Pastor W, Spurney CF, Klugman D, et al. Demographics, trends, and outcomes in pediatric acute myocarditis in the United States, 2006 to 2011. Circulation: Cardiovascular Quality and Outcomes, 2012;5.5: 622–627. doi: 10.1161/CIRCOUTCOMES.112.965749.

21. Butts RJ, Boyle GJ, Deshpande SR, Gambetta K, Knecht KR, Prada-Ruiz CA. Characteristics of clinically diagnosed pediatric myocarditis in a contemporary multi-center cohort. Pediatric cardiology, 2017;38: 1175–1182. doi: 10.1007/s00246-017-1638-1.

22. Park JH, Byeon HJ, Lee KH, Lee JW, Kronbichler A, Eisenhut M, et al. Delta neutrophil index (DNI) as a novel diagnostic and prognostic marker of infection: a systematic review and meta-analysis. Inflammation Research, 2017;66: 863–870. doi: 10.1007/s00011-017-1066-y.

23. McMurray JC, May JW, Cunningham MW, Jones OY. Multisystem inflammatory syndrome in children (MIS-C), a post-viral myocarditis and systemic vasculitis—a critical review of its pathogenesis and treatment. Frontiers in pediatrics, 2020;8: 626182. doi: 10.3389/fped.2020.626182.

24. Foerster SR, Canter CE, Cinar A, Sleeper LA, Webber SA, Pahl E, et al. Ventricular remodeling and survival are more favorable for myocarditis than for idiopathic dilated cardiomyopathy in childhood: an outcomes study from the Pediatric Cardiomyopathy Registry. Circulation: Heart Failure, 2010;3.6: 689–697. doi: 10.1161/CIRCHEARTFAILURE.109.902833.

25. Canter CE, Simpson KE. Diagnosis and treatment of myocarditis in children in the current era. Circulation, 2014;129.1: 115–128. doi: 10.1161/CIRCULATIONAHA.113.001372.

26. Bejiqi R, Retkoceri R, Maloku A, Mustafa A, Bejiqi H, Bejiqi R, et al. The diagnostic and clinical approach to pediatric myocarditis: a review of the current literature. Open access Macedonian journal of medical sciences, 2019;7.1: 162. doi: 10.3889/oamjms.2019.010.

27. Suthar D, Dodd DA, Godown J. Identifying non-invasive tools to distinguish acute myocarditis from dilated cardiomyopathy in children. Pediatric Cardiology, 2018;39: 1134–1138. doi: 10.1007/s00246-018-1867-y.

28. Abrar S, Ansari MJ, Mittal M, Kushwaha KP. Predictors of mortality in paediatric myocarditis. Journal of Clinical and Diagnostic Research: JCDR, 2016;10.6: SC12. doi: 10.7860/JCDR/2016/19856.7967.

